# Integration of proteomics and metabolomics – insights into inflammation, metabolic dysregulation, and vascular aspects in AD

**DOI:** 10.1101/2022.02.18.22271208

**Authors:** Kamil Borkowski, Nicholas T. Seyfried, Matthias Arnold, James J. Lah, Allan I. Levey, Chadwick M. Hales, Eric B. Dammer, Colette Blach, Gregory Louie, Rima Kaddurah-Daouk, John W. Newman, Alzheimer’s Disease Metabolomics Consortium

## Abstract

Integration of the omics data, including metabolomics and proteomics, provides a unique opportunity to search for new associations in the context of metabolic disorders, including Alzheimer’ s disease. Using targeted metabolomics, we have previously profiled lipid mediators, including oxylipins, endocannabinoids, bile acids and steroids in 293 CSF and 202 matched plasma samples from AD cases and healthy controls, and identified both central and peripheral metabolites of the inflammation-regulating cytochrome p450/soluble epoxide hydrolase pathway as strong predictors for the AD pathology. Additionally, using proteomics, we have identified five cerebrospinal fluid protein panels, involved in regulation of energy metabolism, vasculature, myelin/oligodendrocyte, glia/inflammation, and synapses/neurons affected in AD, and reflective of AD-related changes in brain. In the current manuscript, using metabolomics-proteomics data integration, we describe new associations between peripheral and central lipid mediators, with the above-described cerebrospinal fluid protein panels. Particularly strong associations were observed between cytochrome p450/soluble epoxide hydrolase metabolites, bile acids and CSF proteins involved in glycolysis, blood coagulation and vascular inflammation and the regulators of extracellular matrix. Those metabolic associations were not observed at the gene-coexpression level in the central nervous system, showing the importance of pathway interaction investigation on the level of the terminal part of the biochemical “ omic” cascade. In summary, this manuscript provides new information regarding the Alzheimer’ s disease, linking both central and peripheral inflammatory cascade of cytochrome p450/soluble epoxide hydrolase and bile acids metabolism with AD-affected processes and illustrates the necessity for the “ omics” data integration to uncover associations beyond gene co-expression.

## Introduction

The development of multiple omics techniques provides a unique opportunity to probe different aspects of metabolic underpinnings of human disorders [1], including Alzheimer’ s disease (AD) [2, 3]. Metabolomics has been shown to be a useful tool in elucidating molecular manifestations of AD pathomechanisms [4, 5]. These include neuroinflammation, a core feature of AD [6], where targeted metabolomic profiling of lipid mediators in matched plasma and cerebrospinal fluid (CSF) samples revealed evidence for a role of soluble epoxide hydrolase (sEH) and ethanolamides (a class of endocannabinoids) in AD-associated pathologies [7]. These metabolomic variables were distributed across the three classes of oxylipins, endocannabinoids (ECs) and bile acids (BAs). Oxylipins are bioactive oxygenated products of polyunsaturated fatty acids (PUFA) that exhibit both pro- and anti-inflammatory actions [8]. Oxylipin biosynthesis involves the actions of cyclooxygenases (COX), lipoxygenases (LOX), cytochrome P450 (CYP), epoxide hydrolases (EH), and reactive oxygen species, and encompass an array of chemical structures including prostaglandins and epoxides, as well as mono-, di- and tri-hydroxylated species [8, 9]. Endocannabinoids (ECs), named for the ability to activate the CB1 and CB2 cannabinoid receptors, are mainly fatty acid esters and amides that are implicated in regulation of both energy metabolism [10] and inflammatory processes [11]. However, the physiological impacts of ECs are not solely dependent on the CB1 and CB2 activation. ECs and like-substances also interact with the transient potential vanilloid receptor subfamily V member 1 (TRPV1), the G-protein-coupled receptor GPR55 [10] and the peroxisome proliferator-activated receptors (PPARs) [12]. Each of these EC-sensitive receptors are highly expressed in the central nervous system (CNS) [13]. Primary BAs are generated by the liver and secreted into the gut to aid lipid digestion, where they are transformed by the gut microbiome into secondary BAs [14]. After reabsorption from the gut into the blood stream, BAs regulate energy homeostasis with different potency between primary and secondary species mediated through interactions with the farnesoid X receptor (FXR) [15] and the G-protein-coupled bile acid receptor TGR5 [16]. Although communication of these metabolites and proteins are well established, their interactions and regulation in the context of AD remain largely elusive.

The proteome and metabolome together are the end products in the biochemical “ omics cascade”, provide a set of quantitative traits that respond to both genetic and environmental factors and constitute the building blocks of biochemical pathways. Because of their complementarity, integrating proteomic with metabolomics features has the potential to reveal molecular dependencies that go beyond genetic or transcriptional regulation. Previously, we described five functional protein panels in CSF, representing distinct physiological processes that are reflective of AD-associated changes in the brain [17]. These panels include a synaptic panel of neuronal proteins, a vascular panel of endothelial proteins involved in blood coagulation and interaction with extracellular matrix, a myelination panel of oligodendrocyte markers and cellular proliferation, a glial immunity panel of microglia and astrocyte markers and a metabolic panel of proteins involved in energy regulation and storage (i.e. glycolysis).

Since lipid mediators are the key regulators of inflammation among an array of other physiological processes, characterization of how specific peripheral and CSF lipid mediators interact and aggregate with the described functional protein domains in the context of neuroinflammatory processes in AD has the potential to enhance our mechanistic understanding of disease pathology. Alzheimer’ s disease metabolomics consortium (ADMC) is a part of accelerated medicine partnership initiative (AMP) and provides a vast repository of the omics data collected from multiple AD-related cohorts. In the current study, we consequently used ADMC repository of a matched dataset of CSF proteomic panels with CSF and plasma lipid mediator panels in a cohort of participants with and without AD, to probe interactions between these neighboring levels of the omics cascade. We report new interactions between peripheral and central CYP/sEH and BAs metabolism and energy and vascular metabolism in the CNS, describing new connection of inflammation and liver/gut microbiome metabolism and AD.

## Materials and methods

### Subjects

This manuscript contains a secondary analysis of previously described studies [7, 17] with demographics previously summarized [7]. All participants from whom plasma and CSF samples were collected provided informed consent under protocols approved by the Institutional Review Board at Emory University. All protocols were reviewed and approved by the Emory University Institutional Review Board. Cohorts included the Emory Healthy Brain Study (IRB00080300), Cognitive Neurology Research (IRB00078273), and Memory @ Emory (IRB00079069). All patients received standardized cognitive assessments (including Montreal Cognitive Assessment (MoCA)) in the Emory Cognitive Neurology clinic, the ADRC and affiliated Emory Healthy Brain Study (EHBS) [18]. All diagnostic data were supplied by the ADRC and the Emory Cognitive Neurology Program. CSF was collected by lumbar puncture and banked according to 2014 ADC/NIA best practices guidelines. All CSF samples collected from research participants in the ADRC, Emory Healthy Brain Study, and Cognitive Neurology clinic were assayed for total Tau, phosphorylated Tau and AB42 using the INNO-BIA AlzBio3 Luminex assay at AKESOgen (Peachtree Corners, GA). AD cases and healthy individuals were defined using established biomarker cutoff criteria for AD for each assay platform [19, 20]. In total, this analysis utilized data from 202 fasting plasma samples (60 AD cases and 142 healthy controls) and 293 CSF samples (151 AD cases and 142 healthy controls). Two hundred two plasma and CSF samples were matched and collected at the same day.

### Quantification of lipid mediators

Lipid mediators from multiple functional domains and metabolite classes were quantified in 202 plasma and 293 CSF using internal standard methodologies and liquid chromatography tandem mass spectrometry (LC-MS/MS) as reported previously [7, 21]. Briefly, concentrations of non-esterified PUFA, oxylipins, endocannabinoids, a suite of conjugated and unconjugated BAs, and a series of glucocorticoids, progestins and testosterone were quantified by liquid chromatography tandem mass spectrometry (LC-MS/MS) after protein precipitation in the presence of deuterated metabolite analogs (i.e. analytical surrogates). Quality control measures included case/control randomization, and the analysis of batch blanks, pooled matrix replicates and NIST Standard Reference Material 1950 – Metabolites in Human Plasma (Sigma-Aldrich, St Louis, MO) (two per experimental batch). Extracted samples were re-randomized within batch for acquisition, with method blanks and reference materials and calibration solutions scattered regularly throughout the set.

### Quantitative CSF Proteomics

Previously published tandem mass tag (TMT) RAW data of LCMS/MS tryptic peptide digests of whole CSF from 293 individuals were analyzed using the Proteome Discoverer Suite (version 2.3, ThermoFisher Scientific) [17, 22]. Briefly, MS/MS spectra were searched against the UniProtKB human proteome database (downloaded April 2015 with 90,411 total sequences). The Sequest HT search engine was used to search the RAW files, with search parameters specified as follows: fully tryptic specificity, maximum of two missed cleavages, minimum peptide length of 6, fixed modifications for TMT tags on lysine residues and peptide N-termini (+229.162932 Da) and carbamidomethylation of cysteine residues (+57.02146 Da), variable modifications for oxidation of methionine residues (+15.99492 Da), serine, threonine and tyrosine phosphorylation (+79.966 Da) and deamidation of asparagine and glutamine (+0.984 Da), precursor mass tolerance of 20 ppm, and a fragment mass tolerance of 0.6 Da. Percolator was used to filter PSMs and peptides to an FDR of less than 1%. Following spectral assignment, peptides were assembled into proteins and were further filtered based on the combined probabilities of their constituent peptides to a final FDR of 1%. In cases of redundancy, shared peptides were assigned to the protein sequence in adherence with the principles of parsimony as implemented in the Proteome Discoverer software. Reporter ions were quantified using an integration tolerance of 20 ppm with the most confident centroid setting. We have previously described identification of 46 CSF proteins that corresponds to AD-related changes in brain [17]. Five of those proteins were exclude from the analysis in this manuscript due to >25% of missing variables. Remining 41 proteins were converted into the zscores and subsequently into residuals of age, sex, race and ApoE genotype effects, to account for confounders. Protein classification according to GO annotations as well as more in-depth functional classification based on the current literature is presented in the **Table S1**.

### Metabolite – Protein interactions

To explore connections between CNS metabolites and protein expression, we examined correlations between 41 CSF proteins affected by AD with 40 CSF lipid mediators and important metabolite ratios. Associations were assessed using Spearman’ s rank order correlation to account for non-linear associations, using JMP software (SAS institute, Cary, NC). Similarly, to investigate associations between plasma metabolites and CSF proteins, we calculated correlations between the 41 CSF proteins with 93 detected plasma metabolites, including oxylipins, endocannabinoids and fatty acids and their important ratios. Multiple comparison control was accomplished with the false discovery rate (FDR) correction method of Benjamini and Hochberg with a q =0.2 [23]. These analyses were performed separately for participants with and without diagnosed AD, allowing an investigation of the effect of the disease on those associations. Additionally, full factorial linear model was used to test for the metabolite x disease state interaction. Prior to analysis, we converted all variables into residuals of age, sex, race and ApoE genotype effects, to account for confounders. Additionally, we used variable clustering to further illustrate the associations between CSF and plasma metabolites and CSF proteins. To this end, we used an implementation of the PCA based VARCLUS algorithm (JMP, SAS institute, Cary, NC) to cluster metabolites and proteins showing the greatest number and strongest interactions, such as sEH metabolites and proteins highlighted in **Figures 1** and **4**.

**Figure 1.**
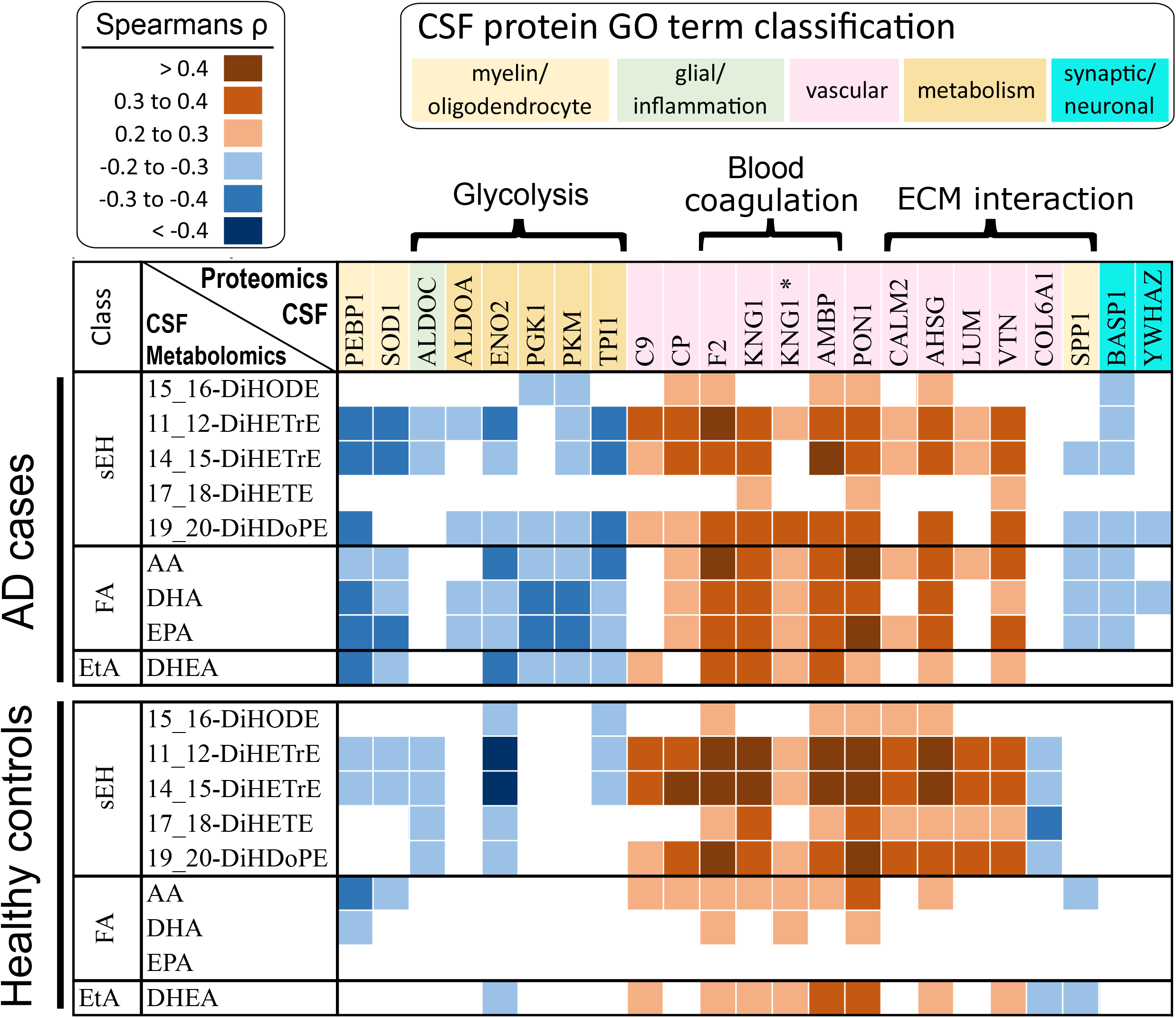
Spearman’ s *ρ* correlations between AD-implicated CSF proteins and CSF metabolites, including oxylipins, fatty acids and endocannabinoids. Only significant associations (p <0,05 and passing FDR corrected at q =0.2) are displayed. Associations are colored according to the Spearman’ s ρ, with positive associations on red and negative on blue scale. Proteins are colored according to Gene Ontology (GO) annotations. Additionally, proteins involved in glycolysis, blood coagulation and extracellular matrix (ECM) interaction are indicated. Spearman’ s *ρ* correlations between all analyzed CSF proteins and CSF metabolites are presented in the **Table S3**. N for AD cases = 151; healthy controls = 142. Two splicing variants are reported for KNG1 (P01042 and P01042-2, marked with asterisk).

**Figure 2.**
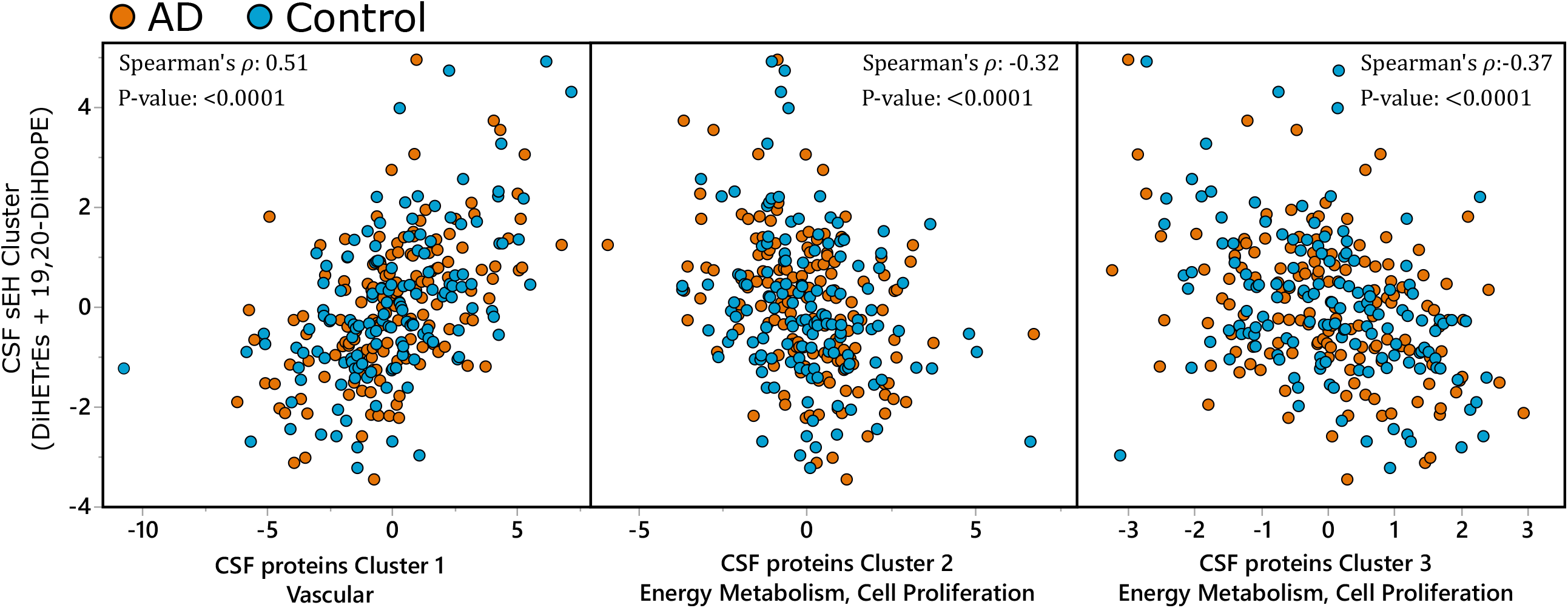
CSF sEH metabolite associations with CSF protein clusters. Most pronounced associations from Figure 1 collapsed into variable cluster components. The sEH metabolite cluster was created from 11,12-DiHETrE, 14,15-DiHETrE and 19,20-DiHDoPE. CSF proteins were organized into three clusters (**Table S5**): Cluster 1 – containing mostly vascular proteins, with F2, AMBP and VTN as most representative members with R^2^>0.8; Cluster 2 – containing proteins involved in energy metabolism and cell proliferation, with PKM, ALDOA and PEBP1 as most representative members with R^2^>0.7; Cluster 3 containing glycolysis regulating ENO2 and cell proliferation regulating COL6A1 and glycolytic enzyme ALDOC. Analysis was performed together for AD cases and healthy controls and the two groups are indicated on the graph by colors: blue – healthy control; orange – AD cases. N for AD cases = 151; healthy controls = 142.

**Figure 3.**
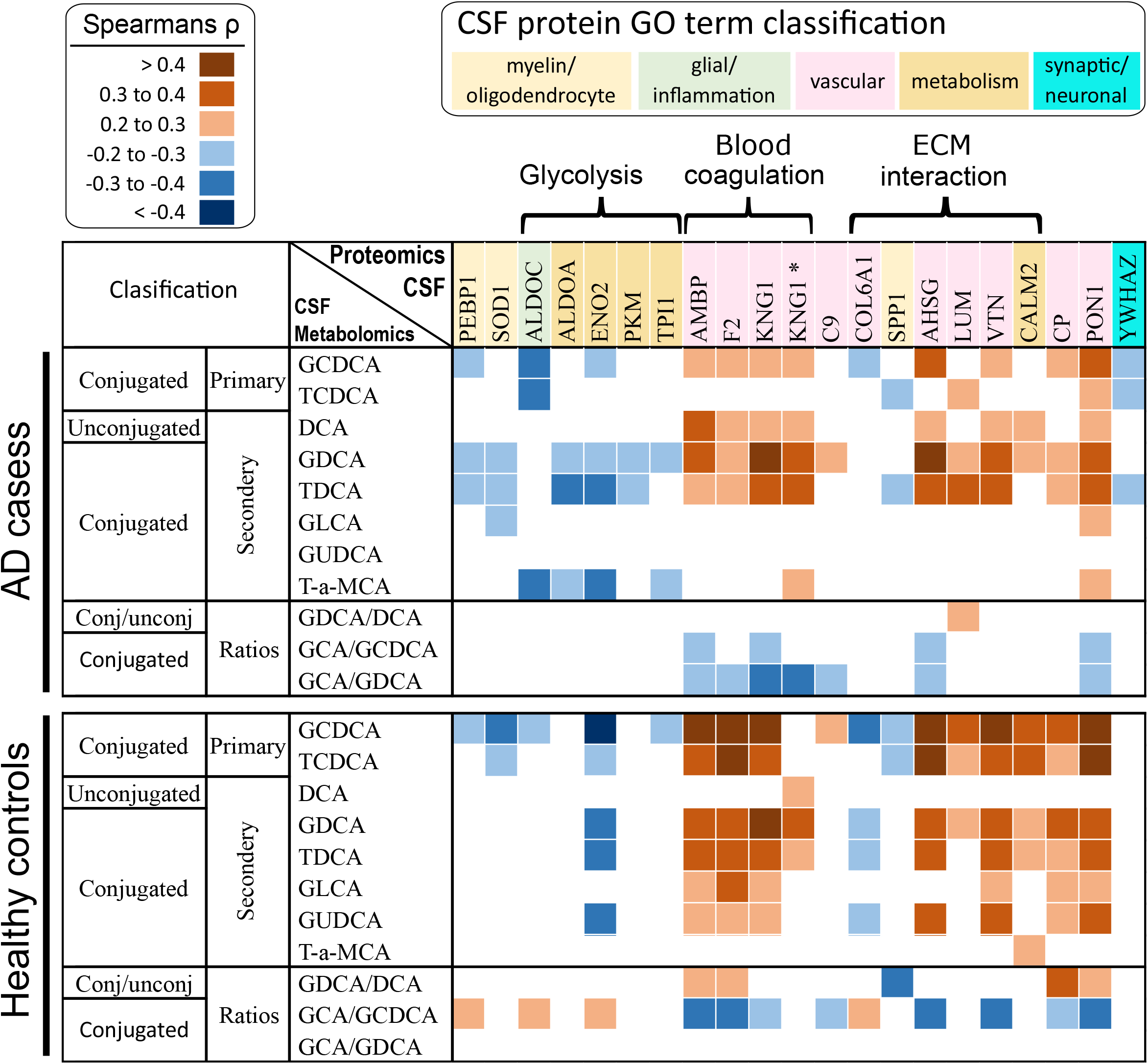
Spearman’ s *ρ* correlations between AD-implicated CSF proteins and CSF BAs. Only significant associations (p<0,05 and passing FDR corrected at q=0.2) are displayed. Associations are colored according to the Spearman’ s ρ, with positive associations on red and negative on blue scale. Proteins are colored according to Gene Ontology (GO) annotations. Additionally, proteins involved in glycolysis, blood coagulation and extracellular matrix (ECM) interaction are indicated. Spearman’ s *ρ* correlations between all analyzed CSF proteins and CSF metabolites are presented in **Table S3**. N for AD cases = 151; healthy controls = 142. Two splicing variants are reported for KNG1 (P01042 and P01042-2, marked with asterisk).

**Figure 4.**
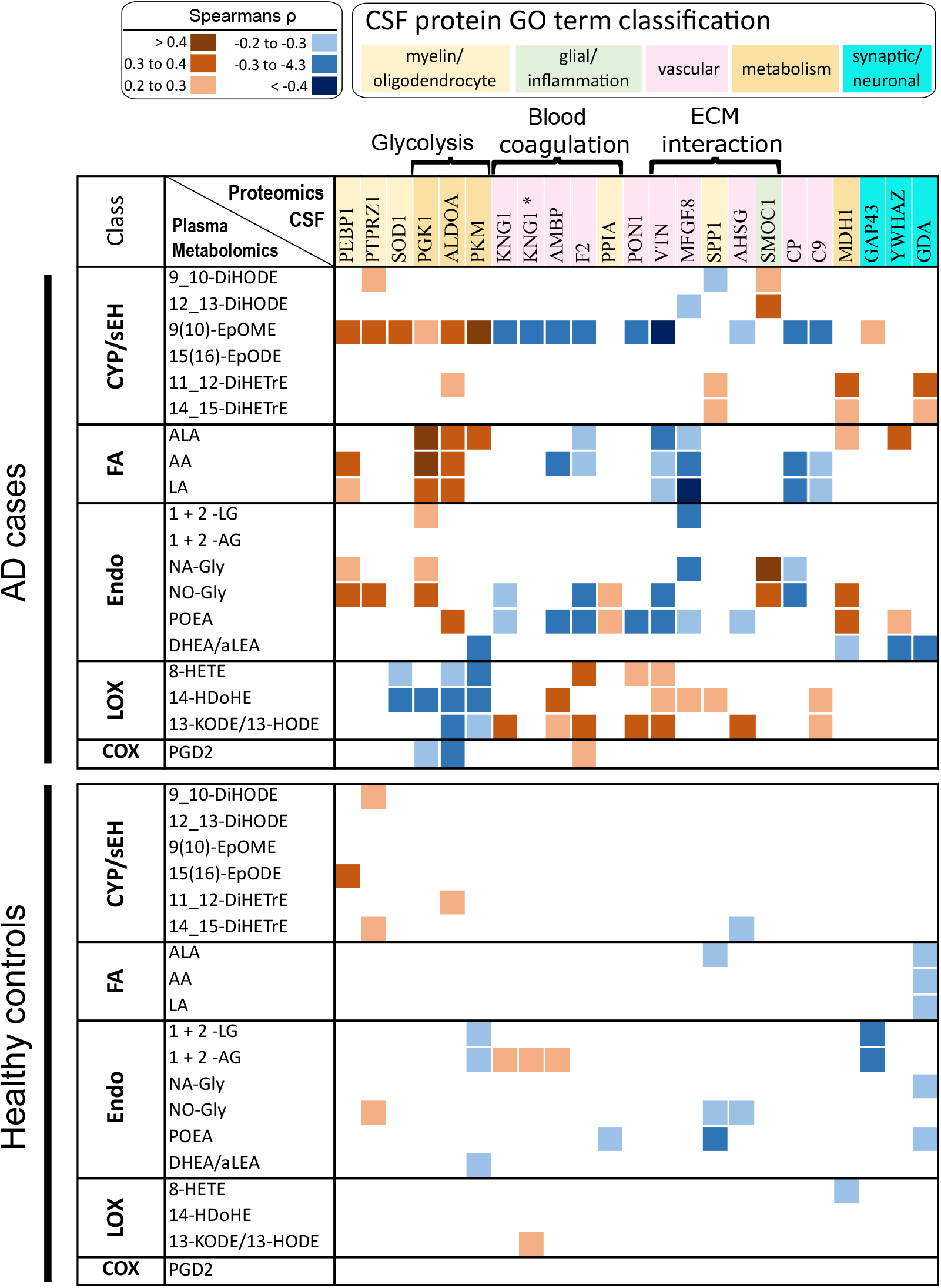
Spearman’ s *ρ* correlations between AD-implicated CSF proteins and plasma metabolites, including oxylipins, fatty acids and endocannabinoids. Only significant associations (p<0,05 and passing FDR corrected at q=0.2) are displayed. Associations are colored according to the Spearman’ s ρ, with positive associations on red and negative on blue scale. Proteins are colored according to Gene Ontology (GO) annotations. Additionally, proteins involved in glycolysis, blood coagulation and extracellular matrix (ECM) interaction are indicated. Spearman’ s *ρ* correlations between all analyzed CSF proteins and plasma metabolites are presented in the **Table S6**. N for AD cases = 60; healthy controls = 142. Two splicing variants are reported for KNG1 (P01042 and P01042-2, marked with asterisk).

**Figure 5.**
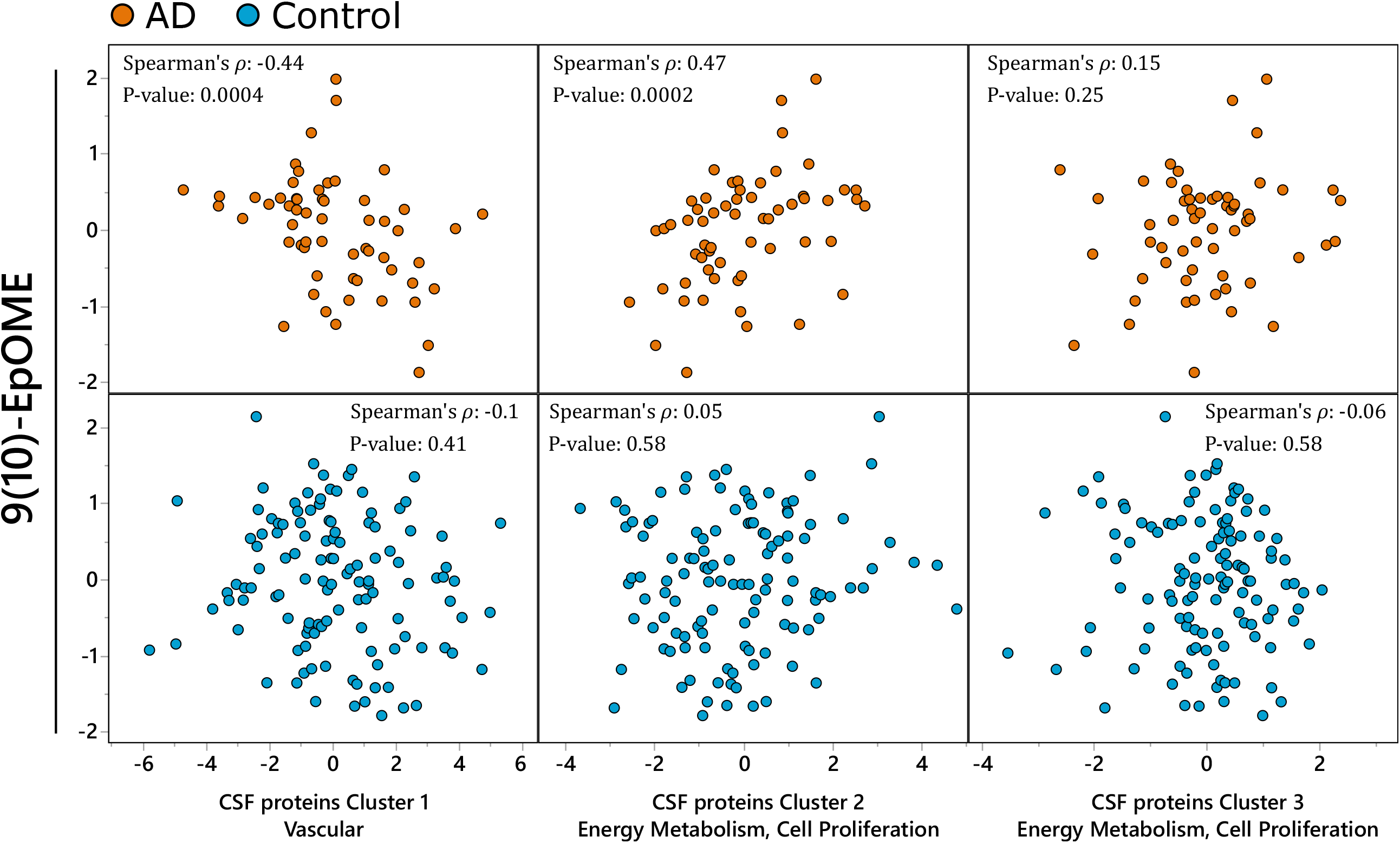
Plasma CYP associations with CSF proteome. Most representative associations from Figure 4, including plasma CYP LA metabolite – 9(10)-EpOME and CSF proteins collapsed into three cluster components (**Table S5)**: Cluster 1 – containing mostly vascular proteins, with F2, AMBP and VTN as most representative members with R^2^>0.7; Cluster 2 – containing proteins involved in energy metabolism and cell proliferation, with PKM, ALDOA and PEBP1 as most representative members with R^2^>0.7; Cluster 3 containing glycolytic enzyme PGK1 and cell proliferation regulating SMOC1 and PPIA. Analysis was performed separately for AD cases and healthy controls due to significant difference in observed associations. N for AD cases = 60; healthy controls = 142.

**Figure 6.**
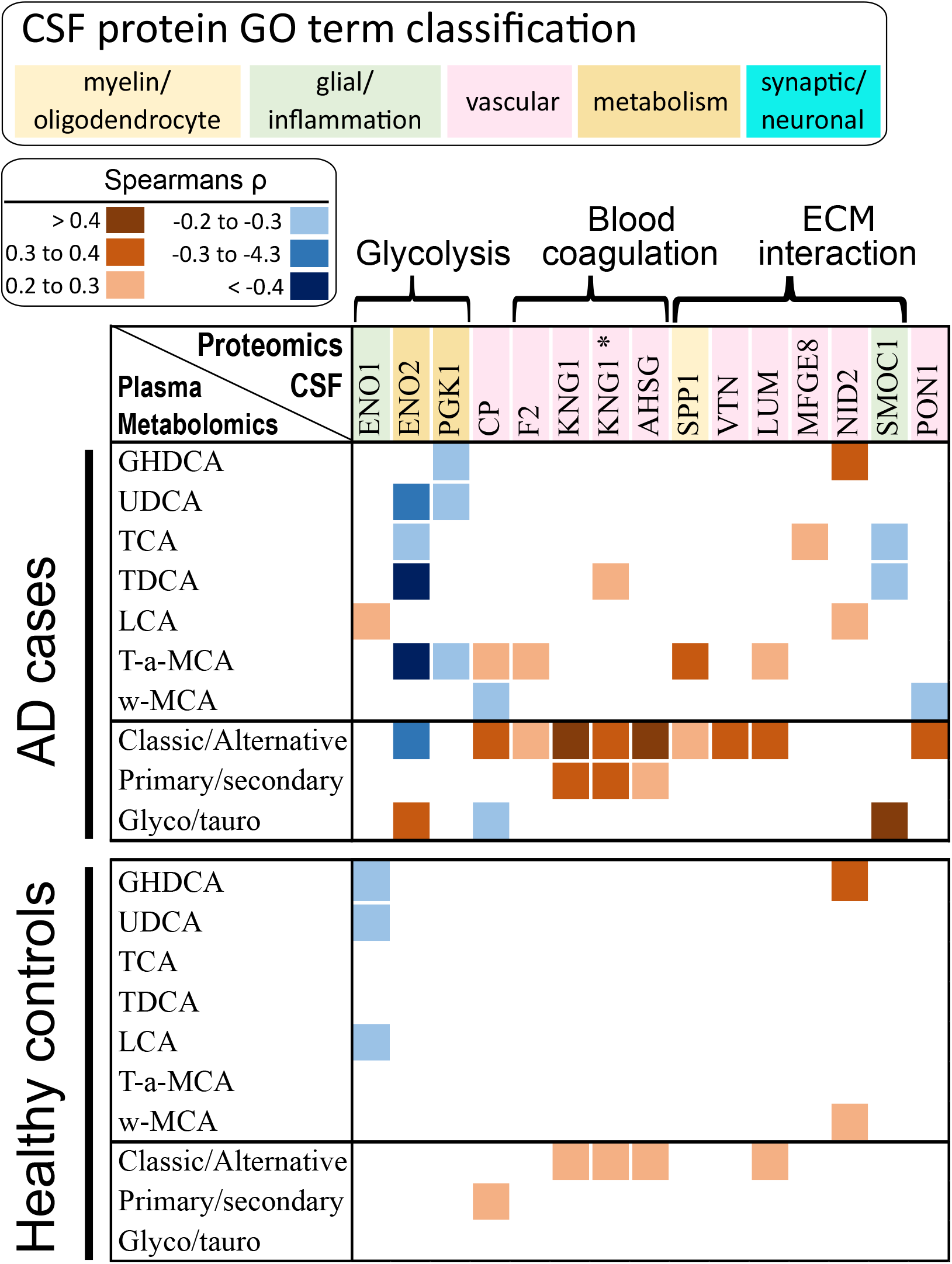
Spearman’ s *ρ* correlations between AD-implicated CSF proteins and plasma BAs. Only significant associations (p<0,05 and passing FDR corrected at q=0.2) are displayed. Proteins are colored according to Gene Ontology (GO) annotations. Additionally, proteins involved in glycolysis, blood coagulation and extracellular matrix (ECM) interaction are indicated. Spearman’ s *ρ* correlations between all analyzed CSF proteins and plasma metabolites are presented in the **Table S6**. N for AD cases = 60; healthy controls = 142. Two splicing variants are reported for KNG1 (P01042 and P01042-2, marked with asterisk). Classic/Alternative pathway is assessed using a ratio of conjugated BAs (TCA+GCA+TDCA+GDCA)/(GUDCA+TUDCA+GLCA+TLCA+TCDCA+GCDCA); Primary/Secondary ratio was assessed using DCA/CA ratio; Glyco/Tauro ratio was assessed using (GDCA+GLCA)/(TDCA+TLCA).

**Figure 7.**
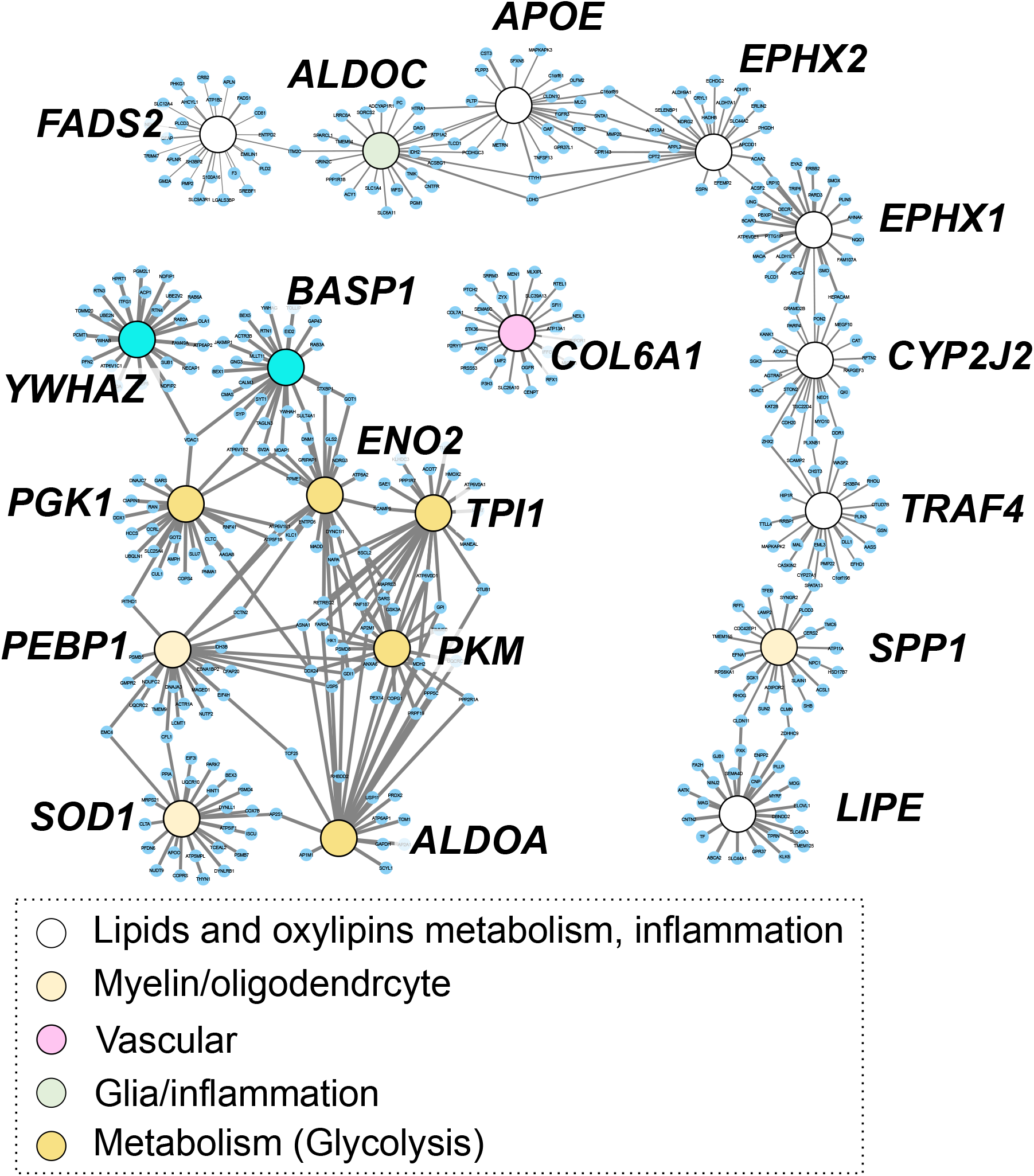
Cerebral gene co-expression network for AD-affected proteins and genes involved in oxylipins and lipids metabolism and inflammation. Nodes are colored according to gene annotation, analogical to Figures 1-4. White nodes represent genes involved in: oxylipins metabolism like main isoform of cytochrome P450 epoxygenase (CYP2J2), soluble epoxide hydrolase (EPHX2) microsomal epoxide hydrolase (EPHX1); lipid metabolism including fatty acids desaturase 2 (FADS2), apolipoprotein E (ApoE), hormone sensitive lipase (LIPE); inflammation - TNF Receptor Associated Factor 4 (TRAF4).

**Figure 8.**
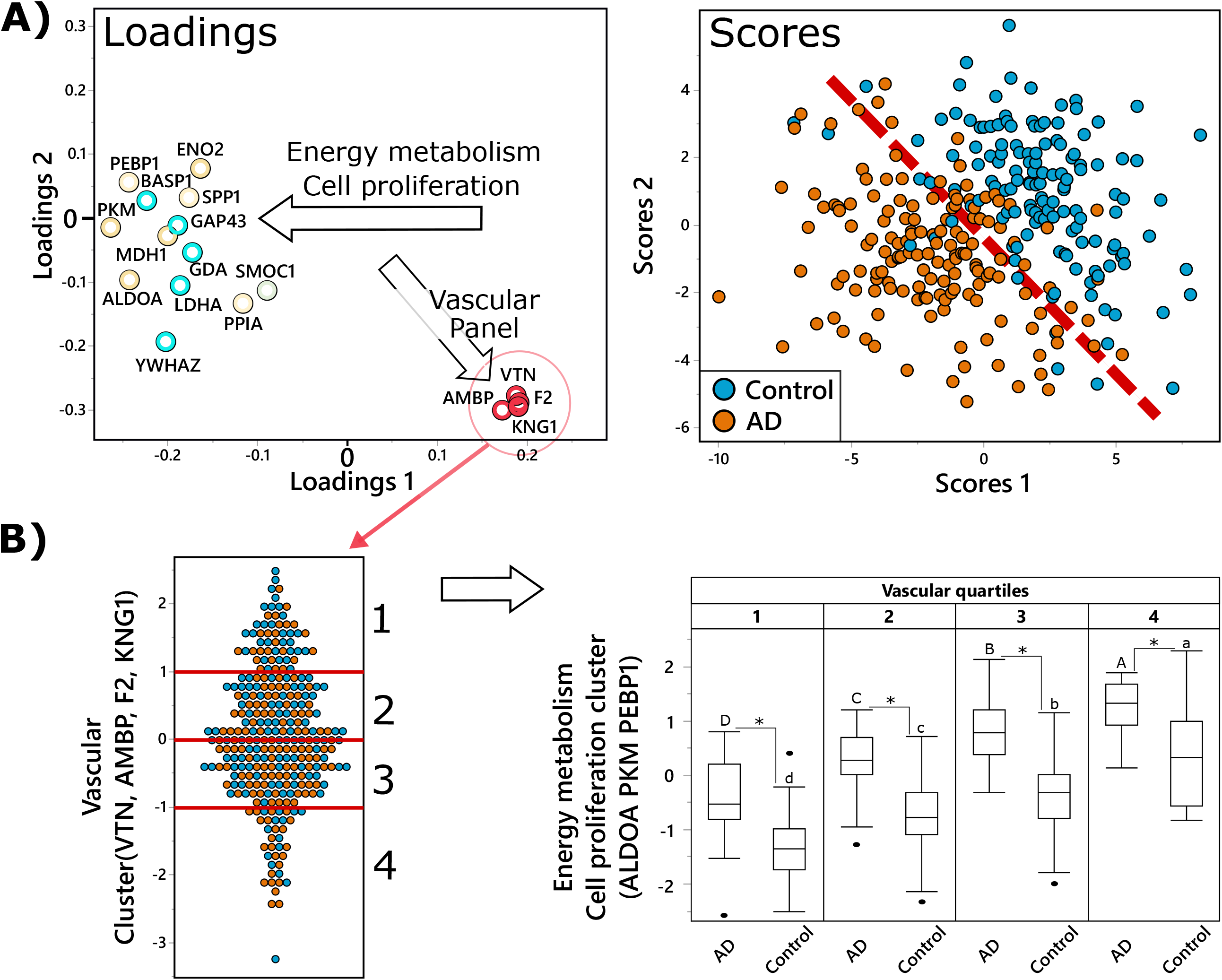
Vascular panel proteins modify relation between energy metabolism-related proteins and AD. A) Partial least square discriminant analysis (PLS-DA) of AD (n=151) vs control (n=142), utilizing CSF proteins. Treatment group discrimination is shown by the SCORES (right panel) with a plane of discrimination indicated by dashed read line, while metabolites weighting in group discrimination are shown by the LOADINGS (left panel). Proteins in the loading plot are colored based on functional panels, same as in in **Figure 1-4**. To facilitate interpretation, arrows on the loading plot indicate directionality, arrow labels indicate the function of discriminating proteins. Analysis was performed with all measured proteins, but only those with variable importance in projection (VIP) ≥1 are displayed for clarity. **B)** Illustration of the relationship between vascular panel proteins (VTN, F2, AMBP, KNG1) and key proteins involved in energy metabolism and cell proliferation (ALDOA, PKM, PEBP1). The panel on the left shows the distribution of the vascular proteins composite score among the subjects, with red lines showing quartiles. The panel on the right shows box and whiskers plots of energy metabolism protein composite score in AD cases and controls, in subjects corresponding to each quartile of the composite score of the vascular panel. Difference in means for each vascular quartile was tested separately for AD cases and healthy controls, using ANOVA with Tukey post-test, with significant differences indicated by capital letters for AD cases and lower letter for the control group. Additionally, differences between AD cases and controls within each vascular quartile was tested using a t-test and significant differences are indicated by an asterisk.

### Cerebral gene co-expression analysis

Potential interaction on a gene expression level between CSF proteins and enzymes involved in oxylipins and fatty acids biology were explored using data from The Genotype-Tissue Expression (GTEx) Project [24] available through the inetmodels.com platform. Network parameters: Tissue – brain cerebrum; tissue type – normal tissue; maximal number of connections pre gene – 25. Additional co-expression data from seven brain regions [25] and genetic associations between input proteins and AD were obtained using the AD Atlas [26]. The list of proteins submitted to the network analyses is provided in **Table S2**.

### Partial least square discriminant analysis

Partial least square discriminant analysis (PLS-DA) was used to investigate the relationship between CSF proteins in discrimination between AD cases and healthy controls, visualizing their multivariate and covariate structure. The PLS-DA model was built using the nonlinear iterative partial least squares algorithm with leave one out cross-validation and included all 41 CSF proteins. For the sake of clarity, we only displayed variables with a variable importance in projection (VIP) score > 1 on the loading plot (Figure 8A).

## Results

### CSF sEH metabolites and PUFA show associations with CSF proteins regulating energy metabolism, vascular function and ECM interaction

Of the 41 tested proteins, 23 showed more than two significant associations with CSF metabolites (**Figure 1**). According to GO classification, 12 of those 23 proteins belong to the vascular panel, five to metabolism panel, three were related to myelin/oligodendrocyte biology, two to synaptic/neuronal function and one to glia/inflammation [17]. Additionally, investigation beyond GO annotations reviled five of those proteins to be involved in glycolysis, four in blood coagulation and six in interaction with the extracellular matrix (ECM) (protein functionality is summarized in the **Table S1**. Spearman’ s ρ and correlation p-values for all associations are presented in **Table S3**.

Soluble epoxide hydrolase (sEH) metabolites, long chain polyunsaturated fatty acids (PUFA) and DHA-derived ethanolamide (DHEA) showed the greatest number of associations. Each metabolite class manifested a similar pattern of associations, including negative correlations with synaptic, neuronal panel, myelin/oligodendrocyte panel and metabolism panel, including six proteins involved in glycolysis, and positive associations with the vascular panel, that included proteins involved in blood coagulation and ECM interactions. Similar patterns of association were observed in individuals with and without diagnosed AD, with some differences. Notably, the healthy control group lacked associations between PUFAs and glycolysis. Additionally, sixteen associations between EPA and CSF proteins showed significant interaction with the disease state in the full factorial linear model, and three interactions were detected for DHA and AA (Interaction p-values for all metabolites are presented in the **Table S4)**. In both groups, the strongest associations were formed with arachidonic acid (AA)-derived 11,12DiHETrE and 14,15-DiHETrE as well as DHA-derived 19,20-DiHDoPE. To summarize and further demonstrate the relations between sEH metabolism and CSF proteins, we analyzed the strongest associations from **Figure 1** using variable cluster components (**Figure 2**). Clusters were formed using CSF sEH metabolites of long chain PUFAS (11,12-DiHETrE, 14,15DiHETrE and 19,20-DiHDoPE, with R square with the cluster = 0.88, 0.87 and 0.51, respectively) and CSF proteins. CSF proteins formed 3 clusters: Cluster 1, represented by vascular proteins F2, AMBP and VTN; Cluster 2 represented by proteins involved in energy metabolism (PKM and ALDOA) and cell proliferation (PEBP1); and Cluster 3 containing the two glycolytic enzymes ENO2 and ALDOC, as well as COL61A, which is involved in ECM synthesis (cluster members and their correlation within each cluster component are described in the **Table S5**). sEH metabolites showed strong positive associations with Cluster 1 (vascular panel), and negative associations with Cluster 2 (cell proliferation and energy metabolism) and cluster 3 (glycolysis, ECM regulation). No interaction with the disease state were detected between sEH metabolites and described CSF protein clusters in the full factorial linear model (P_interaction_= 0.6 for cluster 1, 0.2 for cluster 2 and 1 for cluster 3)

### CSF bile acid associations with CSF proteins

Of the 41 tested proteins, 21 showed more than two significant associations with CSF BAs (**Figure 3**). Eleven of those proteins belong to vascular panel, five to metabolism panel, three to the myelin/oligodendrocyte panel, one to the synaptic/neuronal function panel and one to the glia/inflammation panel. Five of the proteins were involved in glycolysis, four in blood coagulation and six in interaction with ECM. All Bas showed a similar pattern of associations, including negative association with proteins involved in glycolysis and positive associations with vascular panel, that included proteins involved in blood coagulation and ECM interactions (except for COL6A1 and SPP1). Individuals with and without diagnosed AD overall showed similar patterns of associations, however, with some differences. Glycolysis-related proteins were negatively associated only with conjugated BAs, and these were more pronounced among AD case, however, no disease state interactions were detected using the linear full factorial model (**Table S4**). Additionally, in both AD cases and healthy controls, proteins involved in blood coagulation and ECM interaction showed strong and mostly positive associations with the conjugated secondary BAs GDCA and TDCA. Additionally, healthy controls manifested strong positive associations with conjugated primary BAs – GCDCA and TCDCA. Those associations were less pronounced among AD cases and five of those proteins manifested significant disease state interactions in the full factorial linear model. The Spearman’ s ρ and correlation p-values for all associations are presented in **Table S3**.

### Plasma CYP metabolism is associated with CSF proteins regulating energy metabolism, vascular function, and ECM interaction

Of 41 tested proteins, 23 showed more than two significant associations with plasma metabolites (**Figure 4**). Ten of those proteins belong to vascular panel, four to the metabolism panel, five to the myelin/oligodendrocyte panel, three to synaptic/neuronal function panel and one to the glia/inflammation panel. Three of the proteins were involved in glycolysis, five in blood coagulation and five in interaction with ECM.

Of the metabolite classes investigated, the greatest number of associations were observed with cytochrome p450/sEH metabolites, PUFA, endocannabinoids, several hydroxy-fatty acids and prostaglandin D2 (PGD2). Individuals with and without diagnosed AD showed distinct metabolite-protein correlations. In AD cases, the strongest associations were observed between the CYP-derived LA epoxide 9(10)-EpOME, and included positive associations with glycolysis proteins, negative associations with blood coagulating proteins, and strong but mixed associations with ECM interaction proteins. Similarly, three of the five measured PUFAs, aLA, LA and AA showed positive associations with two glycolysis proteins (PGK1 and ALDOA) and negative associations with two ECM interaction protein (VTN and MFGE8). Inverse associations were seen with hydroxy fatty acids. The DHA-derived 14-HDoHE and AA-derived 8-HETE and the ratio of LA- derived 13-KODE/13-HEDE and PGD2 showed negative associations with glycolysis proteins and positive associations with blood coagulation proteins and ECM interaction protein. These associations were absent in healthy controls. Spearman’ s ρ and correlation p-values for all associations are presented in **Table S6**. To summarize and further demonstrate the relations between plasma CYP metabolism and CSF proteins, we analyzed the most representative associations shown in **Figure 4** between LA derived CYP metabolite – 9(10)-EpOME and CSF protein using variable cluster components (**Figure 5**). CSF proteins formed three clusters: Cluster 1 represented by vascular proteins F2, AMBP and VTN; Cluster 2 represented by proteins involved in energy metabolism (PKM and ALDOA) and cell proliferation (PEBP1); Cluster 3, containing PGK1 that is involved in energy metabolism, and SMOC1 and PPIA, which are involved in cell proliferation (individual cluster members and their correlation within each cluster components are described in the **Table S5**). The LA epoxide 9(10)-EpOME showed a strong negative association with Cluster 1 (vascular panel), and positive associations with Cluster 2 (cell proliferation and energy metabolism) and no associations with Cluster 3 (glycolysis, cell proliferation) among AD cases. Those associations were not present among healthy controls. Full factorial linear model showed p-values for disease state interaction of 0.09 for Cluster 1, 0.04 for cluster 2 and 0.35 for cluster 3.

### Plasma bile acid associations with CSF proteins

Of the 41 tested proteins, 15 showed more than two significant associations with plasma BAs (**Figure 6**). Ten of those proteins belong to the vascular panel, two to the metabolism panel, one was related to the myelin/oligodendrocyte panel, and two to the glia/inflammation panel. Three of the proteins were involved in glycolysis, four in blood coagulation and six in interaction with ECM. In AD cases, several BAs, including GHDCA, UDCA, TCA, TDCA and T-a-MCA showed negative associations with two glycolytic proteins ENO2 and PGK1. The greatest number of associations were formed with the specific BA ratios representing the relative activity of the classic and alternative BAs synthesis pathways, calculated using conjugated BAs: (TCA+GCA+TDCA+GDCA)/(GUDCA+TUDCA+GLCA+TLCA+TCDCA+GCDCA), that showed negative associations with ENO2 and positive associations with proteins involved in blood coagulation and ECM interaction. Notably, only few of those associations were observed in healthy controls, however, significant disease state interactions were not observed in the linear full factorial model (**Table S4**). Spearman’ s ρ and correlation p-values for all associations are presented in **Table S6**.

### Brain AD-related and CYP/sEH metabolism-related gene co-expression networks

Unlike BAs, lipid metabolism in the CNS is insulated from peripheral metabolism [7]. Therefore, using data from The Genotype-Tissue Expression (GTEx) Project [24] available through inetmodels.com, we explored the co-expression of genes involved in identified lipid mediators and polyunsaturated fatty acid metabolism and proteins affected by AD. All AD-associated proteins presented in the **Figure 1** were submitted to the analysis using the cerebral brain region of healthy subjects as tissue (**Figure 7**). Gene expression data was not available for members of BAs receptors, including the farnesoid X receptor (FXR), pregnane X receptor (PXR), vitamin D receptor (VDR) and G protein-coupled bile acid receptor 1 (TGR5).

Of 23 AD-associated proteins, 12 were present in the GTEx database and all missing proteins belonged to the vascular panel. To interrogate polyunsaturated fatty acids metabolism, we selected genes involved in the PUFA epoxide/epoxide hydrolase pathway. Specific to epoxy fatty acid metabolism we included the only known CYP with PUFA epoxygenase activity in the GTEx database, CYP2J2 [27], the sEH (EPHX2) and the microsomal epoxide hydrolase (EPHX1), recently suggested to play parallel/complimentary roles in epoxy-PUFA degradation [28]. We then extended this to include fatty acid metabolizing enzymes, including hormone sensitive lipase (LIPE) and fatty acid desaturase 2 (FADS2). We then sought to provide context with proteins involved at the intersection of AD and lipid metabolism. To this end, we included apolipoprotein E (ApoE), a critical apolipoprotein involved in PUFA and oxylipin trafficking with clear AD-associations [29] and TNFα Receptor Associated Factor 4 (TRAF4), an inflammatory cascade regulator upregulated in brains of AD cases [30] with expression levels reportedly associated with EPHX2 in cerebrum [31]. Generally, the CSF-AD protein panel formed a tight gene co-expression network distinct from that of lipid metabolizing proteins. Notably, in the lipid/oxylipin metabolism network, ApoE expression was closely associated with that of EPHX2 and ALDOC, while CYP2J2 expression was closely associated with EPHX1 and TRAF4 expression.

We further investigated potential gene co-expression of CSF proteins and above-described lipid mediators -regulating genes using AD Atlas [26] and seven brain regions. Strong associations were observed within investigated proteins, however no associations were observed between those proteins and lipid mediators enzymatic regulators. Additionally, genetic association (p= 1×10^−13^) was observed between EPHX2 gene and AD phenotype.

### Vascular protein panel in CSF changes the dynamics of energy metabolism in AD

To further understand the relationship between different functional CSF protein panels and AD we implemented a partial least square discriminant analysis (PLS-DA) (**Figure 8**). Out of 41 CSF proteins subjected to the analysis, 17 manifested variable importance in projection (VIP) scores >1. Generally, the model performed well with the Q^2^ =0.48 for two factors, achieving good separation between AD and control subjects. AD and control separation was driven by two functional groups of proteins (indicated by arrows in the **Figure 8A**): (1) energy metabolism and cell proliferation proteins (i.e. key glycolytic proteins ALDOA, PKM and cell proliferation regulating PEBP1, all highly correlated in CSF (r^2^ >0.8); (2) vascular panel proteins, involved in blood coagulation and inflammation (VTN, F2, KNG1, AMBP). Noticeable, vascular proteins were not directly discriminating between AD and controls, but rather served as a covariate for the energy metabolism and cell proliferation proteins. To further illustrate the relationship between energy metabolism and vascular function in the context of AD, vascular proteins were converted into a composite score, and subjects were divided into quartiles of the vascular proteins composite score (**Figure 8B, left panel**). Next, we showed that the level of energy metabolism and cell proliferation proteins (converted into one composite score) is higher in the subjects with low levels of vascular proteins composite score (**Figure 8B, right panel**).

## Discussion

In the current manuscript, we have explored associations between peripheral and CSF lipid mediators and bile acid metabolism with an AD-affected CSF proteome. Our analysis identified, to our knowledge for the first time, associations between CYP/sEH-derived metabolites, PUFAs and BAs with proteins involved in energy metabolism and cell proliferation, blood coagulation and vascular inflammation and ECM regulation in the CNS. Notably, both plasma and CSF members of CYP/SEH pathway, fatty acids, and BAs form strong associations with CSF proteins reflective of AD-related changes in brain [17]. Together, these results show the value of the integration of terminal omics data through utilization of previously published datasets.

Accumulating evidence suggests involvement of lipid mediators in AD pathology. Particularly, several oxylipins of the acute inflammation pathway are elevated in AD [32, 33] and compounds which stimulate inflammatory resolution have been suggested for AD treatment [34]. Specific changes in BAs metabolism, including a decrease in primary and an increase in secondary metabolites were also observed in AD subjects [35] and alterations in bile acid metabolizing enzymes were reported in the AD afflicted brain [36]. Notably, some BAs and some steroids manifest neuroprotective functions through activation of steroid receptors [37].

In CSF, AA and DHA products of sEH (DiHETrEs and DiHDoPE) showed positive associations with the vascular protein panel, including proteins involved in inflammation, blood coagulation and ECM interaction (except for COL6A1), and factors involved in immune response (C9) and detoxification (PON1). The involvement of the brain sEH pathway in AD was previously suggested by us [7] and others [38, 39]. CYP/sEH pathway is known to regulate vascular tone [40] and inflammation [41] in the periphery, and the current manuscript further implicates this pathway in regulation of the CNS vascular system, including vascular inflammation and potentially vascular dysfunction in the CNS, a process attributed to AD pathology [42]. Additionally, protein associations with sEH metabolites were similar between healthy controls and AD cases, while associations with PUFAs were more pronounced in clinical AD. The similarity of observed associations in two independent cohorts of AD cases and healthy controls demonstrates reproducibility of findings and suggests potential importance of the CYP/sEH pathway in healthy CNS.

In plasma, we saw strong positive associations between the LA-derived CYP product (9)10-EpOME, and CSF proteins involved in glycolysis, cell proliferation (PEBP1) and protection from free radicals (SOD1) and negative associations with the CSF vascular panel, including blood coagulation and inflammation proteins. Those associations have opposite directionality than in case of CSF sEH products, consistent with the anti-inflammatory nature of CYP products (epoxy fatty acids) and pro-inflammatory nature of their subsequent sEH metabolites (fatty acids diols) [43]. Additionally, described associations between plasma (9)10-EpOME, and CSF proteins were only present in AD patients and not in the healthy controls, suggesting the potential involvement of peripheral metabolism in AD pathology.

Several studies in animal models for AD and humans have implicated the CYP/sEH pathway in pathogenesis of AD (reviewed in [44]), with a recent study demonstrating upregulation of sEH in the brain of AD cases at the gene and protein level [45]. Our own study identified AD related peripheral and central differences in CYP/sEH metabolism [7]. The CYP/sEH pathway is known for its role in regulation of inflammation [46, 47], the cardiovascular system [40], including vascular inflammation [41], ER-stress [48] and mitochondrial dysfunction [49]. While only correlative, the strong associations of both peripheral and central CYP/sEH metabolites with the CSF vascular protein panel, involved in blood coagulation and cell proliferation are intriguing, providing evidence for peripheral processes to regulate CNS and for synergistic regulation of CYP/sEH pathway in both CNS and periphery. Additionally, our results suggest that the differential levels between healthy and diseased energy metabolism/cell proliferation proteins in the CNS depends on the state of CNS vascular system (lower vascular inflammation and blood coagulation corresponds to higher level of energy metabolism and cell proliferation proteins), providing plausibility for indirect interaction of the CYP/sEH pathway with energy and cell proliferation metabolism. There were previous reports linking the CYP/sEH pathway to energy metabolism in the periphery. In particular, sEH deficiency improves glucose homeostasis [50] and kidney insulin sensitivity [51] in mouse models, and a sEH polymorphism is associated with insulin sensitivity in type 2 diabetic subjects [52]. Additionally, sEH inhibition limits mitochondrial damage in a mouse model of ischemic injury [49]. However, to our knowledge, the involvement of the CYP/sEH pathway in CNS energy metabolism was not previously reported. Interestingly, those associations were only observed on metabolite- protein level and not on the gene co-expression level. The disconnect between transcriptomics and proteomics data is well known [53] as the diversity of regulatory mechanism and regulatory factors increases as we move down the “ omics” cascade, further demonstrating the potential and need for integration of metabolomics and proteomics data.

BAs are another group of lipid mediators that showed strong associations with CSF proteins. BAs are known to regulate both inflammation and energy homeostasis through activation of FXR and TGR5 receptors [54], both abundant in the CNS [55]. However, their regulatory function in the CNS is vastly unknown. In our previous work, we have linked brain [36] and plasma [7, 35] BAs levels to AD and cognition. Here we observed negative associations between CSF BAs levels and CSF proteins involved in glycolysis and cell proliferation (PEBP1), protection from free radicals (SOD1) and ECM interaction and positive associations with CSF blood coagulation proteins. These associations were formed by both primary and secondary BAs (mainly CDCA and DCA amino acid conjugates), suggesting involvement of liver metabolism as well as of the gut microbiome. Additionally, AD cases showed similar correlative structures to healthy controls, suggesting functionality of the observed correlations beyond AD pathology. Regulation of glycolysis by BAs, through activation of muscle TGR5 receptor was previously reported in mouse models [56]. Also, positive associations of plasma BAs and blood coagulation markers was reported in humans [57]. However, associations of BAs and CNS energy metabolism and regulators of vascular function, to our knowledge have not been previously reported.

In plasma, correlations between BAs and CSF proteins were carried mostly by the ratio of conjugated BAs generated *via* classic to alternative pathway, suggesting involvement of liver metabolism and by the markers of gut microbiome activity, like the ratio of secondary to primary BAs (DCA/CA). Associations observed here were present only in AD cases and not in healthy controls, suggesting that involvement of peripheral BAs metabolism is dependent on AD case status.

In conclusion, this study describes new connection between peripheral and central CYP/sEH and BAs metabolism and CNS energy metabolism, cell proliferation, and vascular function. Additionally, our work highlights the potential of multi-omics data integration and shows the need of further cohort analysis in a multi-omics fashion with matching samples, to enable a more in-depth molecular understanding of AD-associated metabolic perturbations.

## Data Availability

Metabolomics data is provided by the Alzheimer's Disease Metabolomics Consortium (ADMC). Metabolomics data and pre-processed data are accessible through the Accelerating Medicines Partnership for AD (AMP-AD) Knowledge Portal (https://adknowledgeportal.synapse.org/). The AMP-AD Knowledge Portal is the distribution site for data, analysis results, analytical methodology and research tools generated by the AMP-AD Target Discovery and Preclinical Validation Consortium and multiple Consortia and research programs supported by the National Institute on Aging. Proteomic data and additional information on their generation is available from synapse.org at https://www.synapse.org/#!Synapse:syn20821165/wiki/603119.

## Declarations

### Ethics approval and consent to participate

All participants from whom plasma and CSF samples were collected provided informed consent under protocols approved by the Institutional Review Board at Emory University. Cohorts included the Emory Healthy Brain Study (IRB00080300), Cognitive Neurology Research (IRB00078273), and Memory @ Emory (IRB00079069). All protocols were reviewed and approved by the Emory University Institutional Review Board. Ethics approval for metabolomics analysis: Analysis samples shipped from Emory was approved for metabolomic profiling and data analysis by the Institutional Review Board at Duke University (Pro00079616).

### Consent for publication

Not applicable

### Availability of data and materials

Metabolomics data is provided by the Alzheimer’ s Disease Metabolomics Consortium (ADMC). Metabolomics data and pre-processed data are accessible through the Accelerating Medicines Partnership for AD (AMP-AD) Knowledge Portal (https://adknowledgeportal.synapse.org/). The AMP-AD Knowledge Portal is the distribution site for data, analysis results, analytical methodology and research tools generated by the AMPAD Target Discovery and Preclinical Validation Consortium and multiple Consortia and research programs supported by the National Institute on Aging. Proteomic data and additional information on their generation is available from synapse.org at https://www.synapse.org/#!Synapse:syn20821165/wiki/603119.

### Authors’ contributions

NS, JWN, AIL, KB and RD designed the study; KB and MA analyzed data; KB wrote the primary manuscript, MA and JWN provided comprehensive revision of the manuscript; JJL, AIL, CMH, EBD, CB and GL contributed to data collection, experimental design and they reviewed the manuscript.

## Acknowledgements

The investigators within the ADMC, not listed specifically in this publication’ s author’ s list, provided data but did not participate in analysis or writing of this manuscript. A complete listing of ADMC investigators can be found at: https://sites.duke.edu/adnimetab/team/.

